# Susceptibility of Ceftazidime-avibactam in bloodstream infections caused by multidrug-resistant Enterobacterales and Pseudomonas aeruginosa

**DOI:** 10.64898/2026.01.20.26344478

**Authors:** André Ricardo Araujo da Silva, Luisa Benigno Barbosa Araujo da Silva

## Abstract

**Background and objectives:** Ceftazidime-Avibactam (CAZ-AVI) is one of the last options to treat Enterobacteriales and *Pseudomonas aeruginosa* carbapenem-resistant. We aim to describe the susceptibility profile of bloodstream isolates of Enterobacterales and *Pseudomonas aerunosa* to ceftazidime-avibactam (CAZ-AVI) among strains resistant to third- and fourth-generation cephalosporins and/or carbapenems.

**Methods:** We conducted a retrospective descriptive study in two pediatric hospitals of Rio de Janeiro city, Brazil, between January 2023 and February 2025. All blood samples with resistance to third/fourth cephalosporins and/or carbapenem resistance were tested to CAZ-AVI, according to the BRCast methodology. Sensibility of CAZ-AVI and clinical profile of patients and outcomes were described.

**Results:** We analyzed 116 blood samples. Of these, 107/116 (92.2%) were resistant to third/fourth-generation cephalosporins with susceptibility to carbapenems, and 9/116 (7.8%) were resistant to both third/fourth-generation cephalosporins and carbapenems. Overall susceptibility to CAZ-AVI was 107/116 (92.2%). The 116 blood samples represented 73 bloodstream infections (BSI) in 66 patients, including 66 single episodes and 7 persistent BSIs. Of the 73 infections, 69(94.5%) were caused by Enterobacterales and 4 (5.5%) by *Pseudomonas aeruginosa*. Twenty-two (30.1%) infections were detected at hospital admission, and 51 (69.9%) were healthcare-associated infections. Death occurred in 5/73 (6.8%) patients. Length of hospital stay (p=0.01596) were statistically significantly higher in non-survivors compared to survivors. The CAZ-AVI was prescribed for four patients with Enterobacteriales or *Pseudomonas aeruginosa* infections with clearance from the blood.

**Conclusion:** Susceptibility of CAZ-AVI to BSI in children was higher and this antibiotic could be an option to treat carbapenem-resistant infection due to Enterobacteriales and *Pseudomonas aeruginosa*.

## INTRODUCTION

In the last ten years, there has benn a global concern about spread of Carbapenem-resistant Enterobacteriales (CRE) including *Klebsiella pneumoniae* and *Escherichia coli* organisms, and *Pseudomonas aeruginosa and Acinetobacter baumannii*. ^1,2^ The resistance prevalence to carbapenems varies depending on the country and bacterial pathogen studied.

The Global Antimicrobial Resistance and Use Surveillance System (GLASS) initiative of World Health Organization (WHO) have been providing worldwide data on the susceptibility and resistance of these pathogens. For example, in the last 2022 update, the resistance rates to meropenem in blood samples was 60.9% for *Acinetobacter baumannii*, 15.7% for *Klebsiella pneumoniae* and 1.2% for *Escherichia coli*.^3^

Carbapenem-resistance in children usually is lower than adults but historical trends series are also showing increasing in resistant rates. ^4^ In Brazil, the largest country of South America, considering notified cases of Central Line-associated bloodstream infection (CLABSI) in children admitted to pediatric intensive care unit (PICU), data from the Brazilian National Health Surveillance Agency (ANVISA) in 2024, indicated resistance rates of 36.6% for *Pseudomonas aeruginosa*, 30.9% for carbapenems in *Klebsiella pneumoniae* pathogens, 30.2% for *Acinetobacter baumannii*, 10.2% for *Escherichia coli*.^5^

Ceftazidime-avibactam (CAZ-AVI) is a recent treatment option to treat gram-negative infections, particularly those caused by CRE, including *Klebsiella pneumoniae* and *Escherichia coli* and multi-drug resistant *Pseudomonas aeruginosa*. Several reports relates a good profile and safety in different clinical settings such as urinary tract infections (UTI), bacteremia, sepsis and in oncological patients. ^6,7^

Despite recent data on CAZ-AVI use in the pediatric population, there is a lack of information about CAZ-AVI susceptibility and resistance in blood samples, especially those resistant to third and fourth-generation cephalosporins and carbapenems. Our aim is to describe the susceptibility profile of bloodstream isolates of Enterobacterales and *Pseudomonas aeruginosa* to ceftazidime-avibactam (CAZ-AVI) among strains resistant to third- and fourth-generation cephalosporins and/or carbapenems.

## METHODS

A retrospective study was conducted to evaluate the in vitro susceptibility of bloodstream isolates of Enterobacterales and *Pseudomonas aeruginosa* to ceftazidime-avibactam (CAZ/AVI) in pediatric patients (0–18 years) admitted to two pediatric hospitals in Rio de Janeiro, Brazil, between January 2023 and February 2025. The isolates included in the analysis exhibited resistance to third- or fourth-generation cephalosporins and/or carbapenems, as interpreted according to the guidelines of the Brazilian Committee on Antimicrobial Susceptibility Testing (BrCAST). All the samples were tested for CAZ/AVI in automatized equipment (Bactec Fx/BD). Testing for CAZ/AVI is avaliable in our clinical practice since January 2023 and used sistematically for clinical purposes. According to the BrCAST, CAZ-AVI resistance was defined with MIC > 8mg/L for Enterobacteriales and > 4 mg/L for Pseudomonas spp. No exclusion criteria was addopted.

For patient analysis in clinical practice, some individuals had two or more samples collected when an infectious episode was suspected. For the purposes of this study, if a patient had two or more positive samples within 48 hours, it was considered a single episode of infection. If a positive sample persisted beyond the third day after the initial isolation, it was classified as a persistent infection episode. Finally, if a new positive sample was detected after the initial episode, a new infection episode was considered after 14 days.

Infections were considered as related to healthcare assistance if occured after 72h of hospital admission and considered from the community or from other units if blood samples were collected less than 72h of hospital admission. Comorbidity was considerated as all previous disease diagnosed previous the hospital admission and included, but not limited to: cardiopathy, chronical lung diseases, any cancer, metabolic or genetic disease, chronic cutaneous disease.

The hospital A has 130 bed-occupancy with emergency room 24h/7 days, clinical, oncologic and surgical wards and 4 pediatric intensive care units (PICU). The hospital B has 45 bed-occupancy with similar profile of unit A, with exception of absence of surgical and oncologic wards. Both units have maintened an active infection control team operating continously since 2005 and implemented an antimicrobial stewardship program in 2017, which includes CAZ-AVI as a restricted antibiotic available just after prior approval of pediatric infectious disease specialist. CAZ-AVI became available for clinical use in the hospitals in 2020; however microbiological testing was only introduced in 2023.

The data were compiled in an Excel file. We performed a descriptive analysis and used means, medians, Mann-Whitney test for compare continuous variables and Fisher’s exact test for categorical variables. A value of p less than.05 was considered as statistically significant.

The study was submitted and approved by the Ethics Committee of Faculty of Medicine (Universidade Federal Fluminense), under number 5.022.885 dated from October 6, 2021.

## RESULTS

One-hundred eleven six third/fourth cephalosporins resistant and/or carbapenem resistant blood samples were isolated and tested for CAZ/AVI susceptibility. Of them, 107/116 (92.2%) were resistant to third/fourth cephalosporins with carbapenem susceptibility and 9/116 (7.8%) were resistant at the same time to third/fourth cephalosporins and carbapenems. No samples was detected with resistance to carbapenem but with susceptibility to third/fourth cephalosporins. Considering these both groups, only 9/116 (7.8%) samples of both groups presented also resistance to CAZ-AVI, being 7/100 (7%) from third/fourth cephalosporins resistant group and 2/9 (22.3%) from third/fourth cephalosporins + carbapenems resistant group (p=0.0892). The number of resistant samples according to the years studied is shown in the table 1.

**Table 1.** CAZ/AVI profile of resistance according to the year studied and resistance to the third/fourth cephalosporins or third/fourth cephalosporins + carbapenems resistant (Rio de Janeiro 2023-2025)

|  | Only Third /Fourth cephalosporins resistant samples |  | Third /Fourth cephalosporins resistant samples and Carbapenem resistant samples |  |
| --- | --- | --- | --- | --- |
|  | CAZ/AVI susceptible<br>N (%) | CAZ/AVI resistant<br>N (%) | CAZ/AVI susceptible<br>N (%) | CAZ/AVI resistant<br>N (%) |
| **2023** | 29 (27.1) | 0 (0) | 1 (11.1) | 0 (0) |
| **2024** | 66 (61.7) | 7 (6.5) | 5 (55.6) | 2 (22.2) |
| **2025** | 5 (4.7) | 0 (0) | 1 (11.1) | 0 (0) |

The 116 blood samples occurred in 66 patients, being 66 singles episodes and 7 persistent bloodstream infections. The table 2 presented the bloodstream infection according to the year studied.

**Table 2.** Bloodstream infection acccording tho the year studied in patients tested for CAZ-AVI and resistant to third/fourth cephalosporins or third/fourth cephalosporins + carbapenems (Rio de Janeiro, 2023-2025)

| Years | Single episodes of BSI (N) | Persistent episodes of BSI ( N) |
| --- | --- | --- |
| 2025 | 5 | 0 |
| 2024 | 44 | 4 |
| 2023 | 17 | 3 |

**Table 3.** Clinical profile of patiens with bloodstream infections previously resistant to third/fourth cephalosporins or third/fourth cephalosporins and/or carbapenems and tested to CAZ-AVI (Rio de Janeiro jan 2023-fev 2025)

| Variables | Values |
| --- | --- |
| **Gender# N (%)** |  |
| Masculine | 44 ( 66.7) |
| Feminine | 22 (33.3) |
| **Age# mean in months ( range)** | 30 ( 0-208) |
| **Type of infection\* N (%)** |  |
| - Community/From the other units | 22 (30.1) |
| - Healthcare-associated infection | 51 (69.9) |
| **Invasive devices previously infection\* N (%)** |  |
| -Yes | 58 (79.5) |
| -No | 15 (20.5) |
| **Comorbidity# N (%)** |  |
| -Yes | 61 (92.4) |
| - No | 5 (7.6) |
| **Outcomes after 30 days of infection\* N ( %)** |  |
| - Discharged | 48 (65.7) |
| - Still admitted | 19 (26) |
| -Dead | 5 (6.8) |
| -Transferred | 1 (1.5) |

Of 73 infections, 69 (94.5%) were caused by Enterobacteriales and 4 (5.5%) due to *Pseudomonas aeruginosa*. Only seven of 73 infections (9.6%) presented resistance to carbapenems. Twenty infections occurred in 2023, 48 in 2024 and 5 in 2025. Twenty-two (30.1%) infections were detected at hospital admission and 51 (69.9%) were diagnosed after 72h of admission.

We also analysed carbapenem and/or third/fourth cephalosporin generation and lenght of hospital stay before the infection in survivors and not survivors. The patient transferred was not included in this analysis. The results are shown in the table 4.

**Table 4.** Use of carbapenem and/or third/fourth cephalosporin generation and lenght of hospital stay before the infection in survivors and not survivors.

| Variables | Survivors | Not survivors | P value |
| --- | --- | --- | --- |
| Carbapenem and/or third/fourth cephalosporin generation before infection – N/total (%) | 20/61 (32.8) | 4/5 (80) | 0.0547 |
| Length of hospital stay before the infection- Mean in days ( range) | 19.7 ( 0-173) | 39.4 (13-66) | 0.01596 |

The CAZ-AVI was prescribed for four patients, all of whom were infected with Enterobacteriales or *Pseudomonas aeruginosa* resistant to carbapenems. In two of these patients, CAZ-AVI showed susceptibility, and one patient was discharged while the other died. In the remaining two patients, CAZ-AVI was used despite resistance confirmed by the antibiogram; in these cases, one patient died and the other remained hospitalized after 30 days of infection. All four patients experienced clearance of Enterobacteriales or *Pseudomonas aeruginosa* from the blood.

## DISCUSSION

The global fight against antimicrobial resistance includes research, development and strategies to prevent and control agents listed by WHO as critical, such as *Acinetobacter baumannii* and Enterobacteriales carbapenem-resistant and Enterobacteriales third-generation cephalosporin-resistant and high for *Pseudomonas aeruginosa* carbapenem-resistant.^8^

CAZ-AVI is a recent developed broad-spectrum antibiotic with activity against infections caused by drug-resistant Gram-negative bacteria, including AmpC, Extended-spectrum Beta-Lactamase, OXA-48-producing Enterobacterales, *Klebsiella pneumoniae* carbapenemase, and MDR, XDR, ceftazidime-non-susceptible, and carbapenem-resistant P. aeruginosa strains. ^9,10^ Despite its broad and clinically relevant spectrum, CAZ-AVI maintains a favorable profile of sensibility to treat these infections, although increasing of resistance rates have been reported worldwide. A recent meta-analysis on global trends in CAZ-AVI resistance in Gram-negative bacteria confirms this concern.^11^

Antimicrobial resistance in pediatric populations is further complicated by the fact that several agents used to treat third-generation cephalosporin-resistant *Enterobacterales* and carbapenem-resistant *P. aeruginosa*—such as cefiderocol and tigecycline—are either not approved for use in children or are only available in certain countries.^12^ For these reasons, we aimed to evaluate CAZ-AVI susceptibility and resistance in bloodstream infections (BSIs) caused by *Enterobacterales* resistant to third- or fourth-generation cephalosporins and *Enterobacterales* or *P. aeruginosa* resistant to carbapenems. Currently, CAZ-AVI remains one of the last available treatment options for these resistant pathogens.

In our study, CAZ-AVI susceptibility testing in pediatric patients with BSIs due to third- or fourth-generation cephalosporin-resistant *Enterobacterales* and carbapenem-resistant *Enterobacterales* or *P. aeruginosa* showed a favorable profile, with only 7.8% resistance across all tested isolates. These results are consistent with those reported in a global meta-analysis of CAZ-AVI activity against Gram-negative bacteria, although their analysis included multiple infection sites, whereas our data focused exclusively on BSIs.^11^ Data specifically addressing CAZ-AVI susceptibility in pediatric BSIs are scarce; however, a Colombian study including isolates from blood, abdominal fluid, urine, soft tissue, and respiratory tract (both pediatric and adult patients) reported resistance rates of 59.26% for carbapenem-resistant *P. aeruginosa* and 1.64% for MDR *Enterobacterales*. ^13^ Similarly, in a large analysis of the in vitro activity of CAZ-AVI against Gram-negative bacteria in patients with bacteremia and skin/soft-tissue infections, among 29 *K. pneumoniae* isolates from children, 19 were classified as MDR, and 10.53% were non-susceptible to CAZ-AVI—a higher rate than that found in our report.^14^

As expected, at least one comorbidity was present in nearly all children with BSIs in our cohort (92.4%), along with the presence of an invasive device. This finding is similar with a systematic review and meta-analysis of CRE bacteremia in pediatric patients in Latin America and the Caribbean, reported that all 149 analyzed patients had an underlying disease and a central venous catheter.^15^

When survival was analyzed according to prior exposure to carbapenems and/or third- or fourth-generation cephalosporins, no statistically significant association was found. However, length of hospital stay prior to infection was significantly longer among non-survivors. Prolonged hospitalization before infection by non-fermentative Gram-negative bacteria has been reported to last up to 51.1 days, as described in a cohort of 131 children from Turkey. ^16^ In our study, 30-day mortality for all patients was 6.8%, which is lower than the 28% and 8.7% reported in Turkey and China, respectively.^16,17^

In our institution, since CAZ-AVI became available in 2020, its use has remained highly restricted, even in pediatric intensive care units. Both hospitals have experienced infection control teams, and a core component of the local antimicrobial stewardship program is the pre-approval of selected antimicrobials, defined according to local microbiological epidemiology. Under this policy, CAZ-AVI was prescribed to only four patients with resistance profile, all with carbapenem-resistant infections. In all cases, bloodstream clearance was achieved; however, two patients died. In two cases, CAZ-AVI was used as salvage therapy due to resistance identified in initial susceptibility testing. Reports on CAZ-AVI use in pediatric populations remain scarce, with most describing patients with underlying conditions such as malignancy, and infections including urinary tract infection, sepsis, and bacteremia, often in the salvage setting. ^6,7,18^

This study has several limitations. First, although CAZ-AVI has been available since 2020, systematic susceptibility testing was only implemented at the end of 2022; nevertheless, the data presented reflect two years of clinical experience. Second, due to the descriptive design, multivariate logistic regression analyses were not performed to identify potential risk factors for 30-day mortality. Finally, CAZ-AVI prescription patterns in our setting are strongly influenced by an active antimicrobial stewardship program, and our results may be most applicable to institutions with similar characteristics. In conclusion, CAZ-AVI susceptibility in pediatric BSIs was high, suggesting that this antibiotic could be an important therapeutic option for carbapenem-resistant *Enterobacterales* and *Pseudomonas aeruginosa* infections in children.

## Data Availability

All data produced in the present work are contained in the manuscript

## Avaliable of patients data

All data of patients are described in the manuscript

## Authors contribution

Luisa Benigno Barbosa Araujo da Silva-Formal analysis, investigation, writing-review and editing. André Ricardo Araujo da Silva-Conceptualization, formal analysis, investigation, methodology, visualization, writing-original draft prepation, writing-review and editing.

## REFERENCES

1. Ma J, Song X, Li M, Yu Z, Cheng W, Yu Z, Zhang W, Zhang Y, Shen A, Sun H, Li L. Global spread of carbapenem-resistant Enterobacteriaceae: Epidemiological features, resistance mechanisms, detection and therapy. Microbiol Res. 2023 Jan;266:127249. doi: 10.1016/j.micres.2022.127249.

2. Fisher M, Komarow L, Kahn J, Patel G, Revolinski S, Huskins WC, van Duin D, Banerjee R, Fries BC. Carbapenem-resistant Enterobacterales in Children at 18 US Health Care System Study Sites: Clinical and Molecular Epidemiology From a Prospective Multicenter Cohort Study. Open Forum Infect Dis. 2024 Jan 8;11(2):ofad688. doi: 10.1093/ofid/ofad688.

3. Global Antimicrobial Resistance and Use Surveillance System (GLASS). World Health Organization. Available at: https://www.who.int/initiatives/glass. Access on July 22, 2025.

4. Logan LK, Renschler JP, Gandra S, Weinstein RA, Laxminarayan R; Centers for Disease Control; Prevention Epicenters Program. Carbapenem-Resistant Enterobacteriaceae in Children, United States, 1999-2012. Emerg Infect Dis. 2015 Nov;21(11):2014–21. doi: 10.3201/eid2111.150548.

5. Boletins Segurança do Paciente e Qualidade em Serviços de Saúde nº32-Avaliação Nacional dos indicadores de IRAS e RM – 2024. Brazilian National Health Surveillance Agency (ANVISA). Available at: https://www.gov.br/anvisa/pt-br/centraisdeconteudo/publicacoes/servicosdesaude/boletins-e-relatorios-das-notificacoes-de-iras-e-outros-eventos-adversos-1/boletins-e-relatorios-das-notificacoes-de-iras-e-outros-eventos-adversos. Access on July 24, 2025.

6. Zhu L, Hu Q, Liu L, Ye S. Ceftazidime-Avibactam as a Salvage Treatment for Severely Infected Immunosuppressed Children. Drug Des Devel Ther. 2024 Jul 30;18:3399–3413. doi: 10.2147/DDDT.S467967.

7. Castro-Frontiñán A, Prieto-Tato LM, Caro-Teller JM, Epalza C, González-Gómez Á, Villaverde S, Shan-Núñez A, Viedma E, Ferrari-Piquero JM; Hospital 12 de Octubre Pediatric Antimicrobial Stewardship Group. Ceftazidime/Avibactam for the Treatment of Infections in Children: A Case Series of Real-World Use. Paediatr Drugs. 2025 Jul;27(4):427–438. doi: 10.1007/s40272-025-00685-7.

8. WHO bacterial priority pathogens list, 2024: Bacterial pathogens of public health importance to guide research, development and strategies to prevent and control antimicrobial resistance. World Helath Organization, Geneve, 2024. Available at: https://www.who.int/publications/i/item/9789240093461 Acessed on July 27, 2025.

9. Shirley M. Ceftazidime-avibactam: a review in the treatment of serious gram-negative bacterial infections. Drugs. 2018;78:675–92

10. Hobson CA, Pierrat G, Tenaillon O, Bonacorsi S, Bercot B, Jaouen E, et al. Klebsiella pneumoniae carbapenemase variants resistant to ceftazidime-avibactam: an evolutionary overview. Antimicrob Agents Chemother. 2022;66(9):e00447–e522.

11. Wang Y, Sholeh M, Yang L, Shakourzadeh MZ, Beig M, Azizian K. Global trends of ceftazidime-avibactam resistance in gram-negative bacteria: systematic review and meta-analysis. Antimicrob Resist Infect Control. 2025 Feb 11;14(1):10. doi: 10.1186/s13756-025-01518-5.

12. Lockowitz CR, Hsu AJ, Chiotos K, Bio LL, Dassner AM, Gainey AB, Girotto JE, Iacono D, Morrisette T, Stimes G, Tran MT, Wilson WS, Tamma PD. Suggested Dosing of Select Beta-lactam Agents for the Treatment of Antimicrobial-Resistant Gram-Negative Infections in Children. J Pediatric Infect Dis Soc. 2025 Feb 6;14(2):piaf004. doi: 10.1093/jpids/piaf004.

13. Lemos-Luengas EV, Rentería-Valoyes S, Cárdenas-Isaza P, Ramos-Castaneda JA. In vitro activity of ceftazidime/avibactam against Gram-negative strains in Colombia 2014-2018. J Glob Antimicrob Resist. 2022 Jun;29:141–146. doi: 10.1016/j.jgar.2022.02.018.

14. Lemos-Luengas EV, Rentería-Valoyes S, Muñoz DMA, Gonzalez CKG, Guerrón-Gómez G, Ramos-Castaneda JA. In vitro activity of ceftazidime-avibactam against gram-negative bacteria in patients with bacteremia and skin and soft-tissue infections in Colombia 2019-2021. Diagn Microbiol Infect Dis. 2024 Jun;109(2):116235. doi: 10.1016/j.diagmicrobio.2024.116235.

15. Ruvinsky S, Voto C, Roel M, Portillo V, Naranjo Zuñiga G, Ulloa-Gutierrez R, Comandé D, Ciapponi A, Aboud G, Brizuela M, Bardach A. Carbapenem-Resistant Enterobacteriaceae Bacteremia in Pediatric Patients in Latin America and the Caribbean: A Systematic Review and Meta-Analysis. Antibiotics (Basel). 2024 Nov 22;13(12):1117. doi: 10.3390/antibiotics13121117.

16. Guner Ozenen G, Sahbudak Bal Z, Umit Z, Avcu G, Tekin D, Kurugol Z, Cilli F, Ozkinay F. Nosocomial Non-fermentative gram negative bacteria bloodstream infections in children; Risk factors and clinical outcomes of carbapenem resistance. J Infect Chemother. 2021 May;27(5):729–735. doi: 10.1016/j.jiac.2020.12.024.

17. Zhang Y, Guo LY, Song WQ, Wang Y, Dong F, Liu G. Risk factors for carbapenem-resistant K. pneumoniae bloodstream infection and predictors of mortality in Chinese paediatric patients. BMC Infect Dis. 2018 May 31;18(1):248. doi: 10.1186/s12879-018-3160-3.

18. Hoshino WT, da Silva AMPS, Pignatari AC, Gales AC, Carlesse F. Experience in Ceftazidime-Avibactam for treatment of MDR BGN infection in Oncologic Children. Braz J Infect Dis. 2025 Mar-Apr;29(2):104515. doi: 10.1016/j.bjid.2025.104515.

